# SARS-CoV-2 Antibody Response is Associated with Age in Convalescent Outpatients

**DOI:** 10.1101/2021.11.08.21265888

**Authors:** Bo Zhai, Karen Clarke, David L. Bauer, Saran Kupul, Lucas J. Schratz, M. Patricia Nowalk, Anita K. McElroy, James B. McLachlan, Richard K. Zimmerman, John F. Alcorn

## Abstract

COVID-19 has had an unprecedented global impact on human health. Understanding the antibody memory responses to infection is one tool needed to effectively control the pandemic. Among 173 outpatients who had virologically confirmed SARS-CoV-2 infection, we evaluated serum antibody concentrations, microneutralization activity, and enumerated SARS-CoV-2 specific B cells in convalescent blood specimens. Serum antibody concentrations were variable, allowing for stratification of the cohort into high and low responders. Serum antibody concentration was positively associated with microneutralization activity and participant age, with participants under the age of 30 showing the lowest antibody level. Neither participant sex, the timing of blood sampling following the onset of illness, nor the number of SARS-CoV-2 spike protein specific B cells correlated with serum antibody concentration. These data suggest that young adult outpatients did not generate as robust antibody memory, compared with older adults. Further, serum antibody concentration or neutralizing activity trended but did not significantly correlate with the number of SARS-CoV-2 memory B cells. These findings have direct implications for public health policy and current vaccine efforts. Knowledge gained regarding antibody memory following infection will inform the need for vaccination in those previously infected and allow for a better approximation of population-wide protective immunity.

## Introduction

The ongoing COVID-19 pandemic caused by SARS-CoV-2 has resulted in over 200 million cases and 4.5 million deaths worldwide as of October 2021. With a case fatality rate near 2%, most of those infected survive and go on to generate immune memory against the virus. Numerous studies have shown that greater than 95% of those infected generate antibodies that recognize SARS-CoV-2 proteins in the months immediately following infection, as reviewed (1). SARS-CoV-2 infection results in neutralizing antibody production in most of those infected, although the half-life of neutralizing antibody may be a relatively short 2 months (1). Neutralizing antibody production is thought to be protective against re-infection in about 90% of individuals (1). Many studies to date have focused on those hospitalized with severe COVID-19 although several groups have published investigations into the antibody response to SARS-CoV-2 infection in those with less severe disease (2-8). Comparison of the antibody response in patients with severe disease with those with mild/moderate disease or the response induced by vaccination have shown that severe illness results in robust and greater antibody levels than the other settings (9, 10).

Although SARS-CoV-2 infection has been shown to produce a significant antibody response that initially protects against re-infection, it has been shown to wane over the course of months. This waning immune memory suggests a need for immune boosting by vaccination (9). Further, the impact of age and sex on antibody response to SARS-CoV-2 is inconsistent in the literature and may play a key role in defining which patients are most in need of boosting for sustaining adequate immune memory. The goal of the present study was to better understand the antibody response to SARS-CoV-2 infection in adults with mild to moderate illness who sought outpatient care using well characterized and controlled assays for SARS-CoV-2 spike (S1, receptor binding domain (RBD), S2 domains) and nucleoprotein (N) specific IgG antibody. SARS-CoV-2 antibody concentrations were correlated with *in vitro* virus neutralization activity to demonstrate the functional relevance of high and low antibody concentration responders. In addition, the presence of IgG producing memory B cells was examined by flow cytometry in participants with high serum antibody responses to SARS-CoV-2. This detailed examination of the antibody response to SARS-CoV-2 in outpatients was intended to improve the identification of previously infected patients to prioritize for vaccine immune boosting.

## Results

To examine the antibody response to mild SARS-CoV-2 infection, we recruited 173 convalescent outpatients for blood sampling through the FLUVE network. The patient cohort included 105 females with a mean age of 38.3 (range = 19-79) years and 67 males with a mean age of 42.7 (range = 20-78) years. One participant did not disclose their sex. The convalescent blood sample was drawn at an average of 44.5 (range = 22-131) days post symptom onset for females and 42.3 (range = 21-89) days for males. There were no significant differences in age or timing of sampling by participant sex.

SARS-CoV-2 antibodies were detectable in most participant samples. Concentrations ranged from 0 to 2×10^6^ U/ml. Low responders were defined as antibody concentration less than 5 × 10^4^ U/ml. Approximately one-fifth of participants had low antibody levels against N, RBD, and S1 proteins (Table 1). Incidence of low response to S2 was lower than the other antigens tested at 6.9% of participants. Approximately 6% of participants had very low antibody responses (less than 10^4^ U/ml) to N, RBD, and S1, while only a few had very low antibody levels against S2. Of the 18 with the lowest antibody concentrations, 13 were females. Of these 18, low responses were noted against 1 of 4 antigens in 6 persons, 2 antigens in 7 persons, 3 antigens in 3 persons and all 4 antigens in 2 persons.

**Table 1.**

| Antibody | Female (N=105) | Male (N=67) | Total (N=173) |
| --- | --- | --- | --- |
| N |  |  |  |
| <10^4 | 7 (6.7%) | 4 (6.0%) | 11 (6.4%) |
| <5x10^4 | 25 (23.8%) | 13 (19.4%) | 39 (22.5%) |
| RBD |  |  |  |
| <10^4 | 9 (8.6%) | 2 (3.0%) | 11 (6.4%) |
| <5x10^4 | 26 (24.8%) | 12 (17.9%) | 39 (22.5%) |
| S1 |  |  |  |
| <10^4 | 9 (8.6%) | 3 (4.5%) | 12 (6.9%) |
| <5x10^4 | 26 (24.8%) | 12 (17.9%) | 39 (22.5%) |
| S2 |  |  |  |
| <10^4 | 2 (1.9%) | 1 (1.5%) | 3 (1.7%) |
| <5x10^4 | 5 (4.8%) | 6 (9.0%) | 12 (6.9%) |

We examined correlations between antibody levels for pairs of SARS-CoV-2 proteins. The highest correlation (R^2^ = 0.74) in antibody levels was for S1 and RBD, which would be predicted since RBD is part of S1 (Figure 1). Significant correlations between antigen antibody levels were found for all 4 antigens tested. Interesting, N protein antibody levels correlated at the lowest levels in all comparisons. Despite statistically significant correlations, the relationship between individual antigen antibody levels were not overtly linear, as was the case for S1 and RBD.

**Figure 1.**
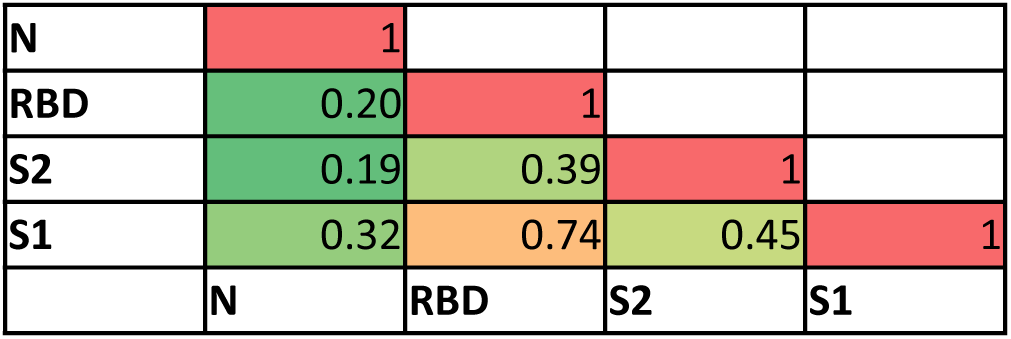
Correlation between SARS-CoV-2 antigen specific IgG concentrations. Samples in the cohort were analyzed for 4 SARS-CoV-2 antigens (N=173). Concentration values (U/ml) were compared using simple linear regression, P ≤ 0.0001 for all comparisons.

To determine if measurable differences in SARS-CoV-2 antibody concentration was relevant to functional antibody, microneutralization assays were performed, on a subset of participants with the most divergent antibody levels. This subset of 56 participants was selected to assess both high and low responders: based upon high antibody response (greater than 7x 10^5^ U/ml for at least 1 antigen) and low antibody response (less than 1 × 10^5^ U/ml for at least 1 antigen). Evidence of neutralizing antibody activity (FRNT_80_) was uncommon in the low responder group with only 9 of 28 participants achieving neutralizing titer ≥ 1:20 (Figure 2). No high titer neutralizing activity was seen in the low responder group (greatest observed 1:80). Conversely, all but 1 high responder displayed neutralizing activity, with 18 of 28 showing a neutralizing titer greater than or equal to 1:160. Further, microneutralization titer significantly correlated with the concentration of antibody determined by Bioplex (Supplemental Figure 1). Correlation was strongest between microneutralization titer and spike protein antigens (e.g., r^2^ of 0.36 for RBD).

**Figure 2.**
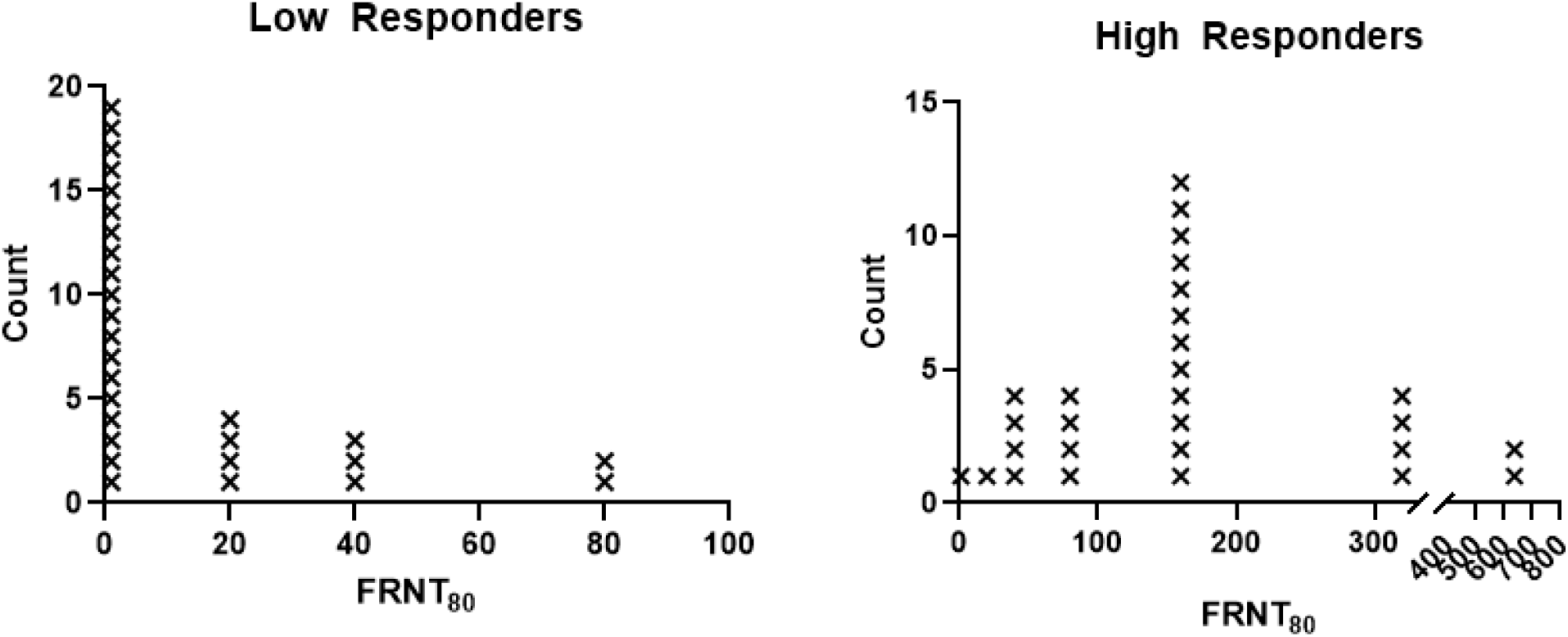
In vitro SARS-CoV-2 microneutralization titers in high and low responding participants. A subset of participants were stratified by relative vaccine response into two groups of lowest and highest antibody concentrations (N=28 each). The dilution of serum at which 80% of viral foci are neutralized is reported as the FRNT_80_ (1:X titer). The limit of detection for the assay is 1:20.

Flow cytometry was then utilized to determine if SARS-CoV-2 specific B, memory B, and IgG producing B cells were detectable in PBMC from high responders. We employed a SARS-CoV-2 spike tetramer and a gating strategy to differentiate total spike-specific B cells, memory B cells, and class switched IgG producing B cells (Supplemental Figure 2). We successfully detected spike specific B cells in all three gates from all high responders. The number of spike-specific B cells trended to positively correlate with microneutralization titer, but not with memory B cell or IgG producing B cell numbers (Figure 3). Spike-specific B cells trended higher in male participants versus female (Figure 4), although no differences were seen in spike-specific memory or IgG producing B cells. Participant age was then compared with the number of spike-specific B cells. SARS-CoV-2 spike-specific memory B cells trended higher in younger participants within the high responder group (Figure 4). There was no correlation between spike-specific B cell numbers and the timing of sampling post-illness or antibody concentration (data not shown).

**Figure 3.**
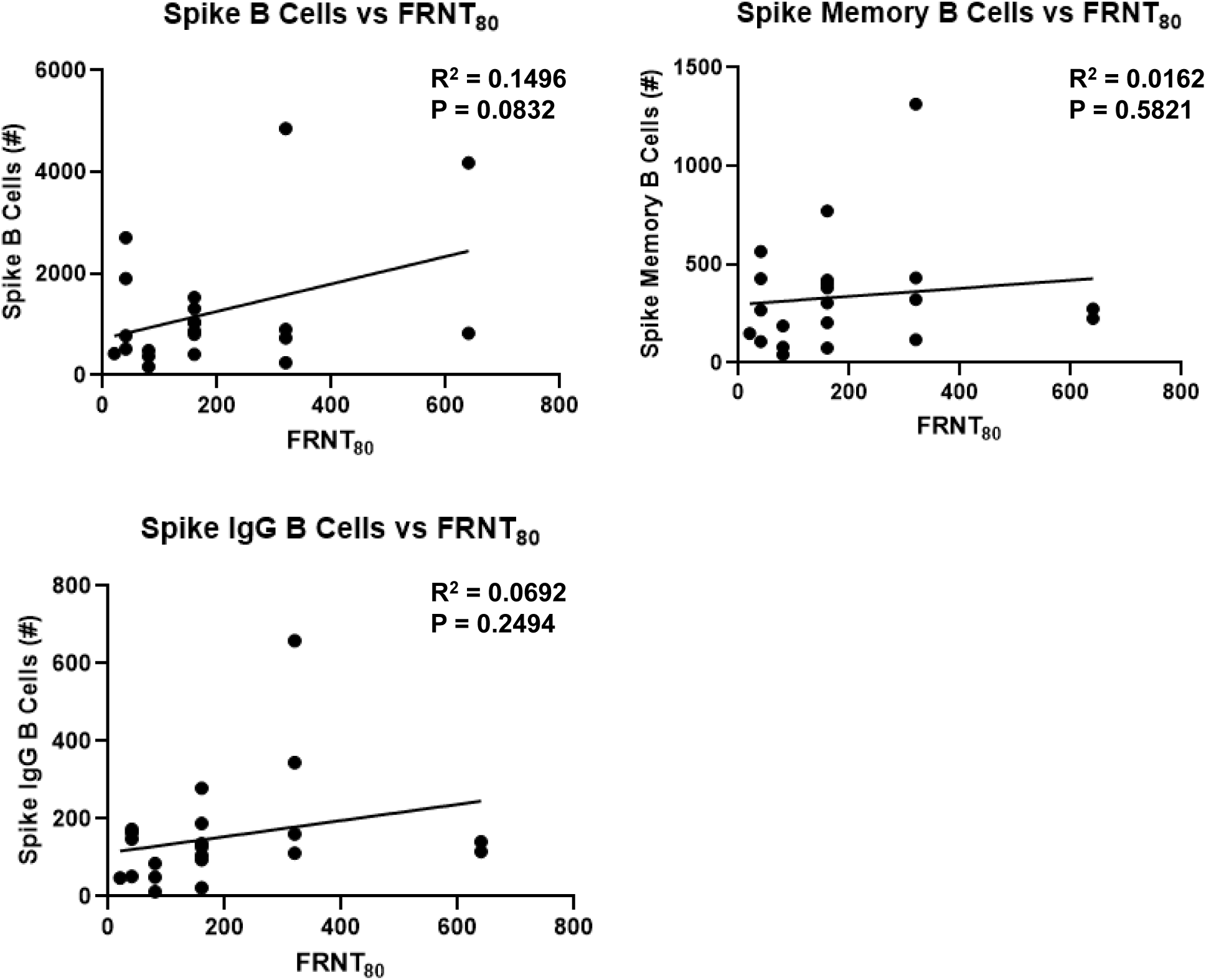
Correlation of spike-specific B cells and serum microneutralization activity. Flow cytometry was performed on high responder participant PBMC using a spike-specific tetramer and B cell markers (N=21). Spike B cells were defined as CD19^+^ tetramer^+^ cells. Spike memory B cells were defined as CD19^+^ tetramer^+^ CD27^+^ cells. Spike-specific IgG B cells were defined as CD19^+^ tetramer^+^ IgD^-^ IgM^-^ IgG^+^ cells. Data were compared using simple linear regression.

**Figure 4.**
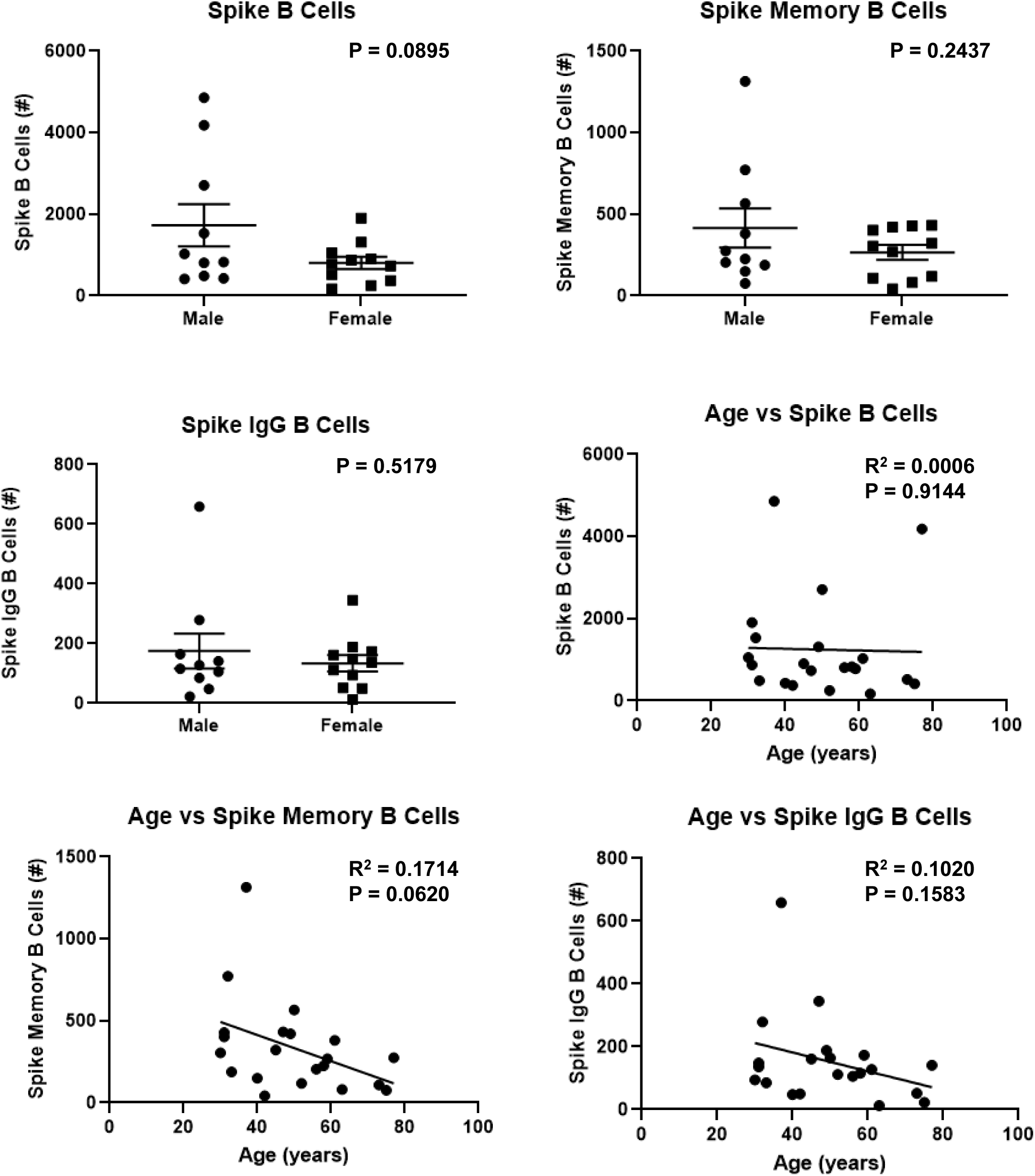
Effect of biologic sex or participant age on SARS-CoV-2 spike specific B memory and IgG producing B cells. Flow cytometry was performed on high responder participant PBMC using a spike-specific tetramer and B cell markers (N=21). Spike B cells were defined as CD19^+^ tetramer^+^ cells. Spike memory B cells were defined as CD19^+^ tetramer^+^ CD27^+^ cells. Spike-specific IgG B cells were defined as CD19^+^ tetramer^+^ IgD^-^ IgM^-^ IgG^+^ cells. Data were compared using unpaired t-test or simple linear regression.

We examined the impact of sample timing and/or biologic variables on SARS-CoV-2 antibody responses in the entire cohort. The mean sampling time was approximately 6 weeks post onset of illness, which was utilized to divide the group into those sampled before and after 42 days post illness onset. There was no significant difference in antibody levels by time of sampling for any of the 4 antigens tested based (Figure 5). Neither was there a difference in antibody concentration against the SARS-CoV-2 antigens tested (Figure 6).

**Figure 5.**
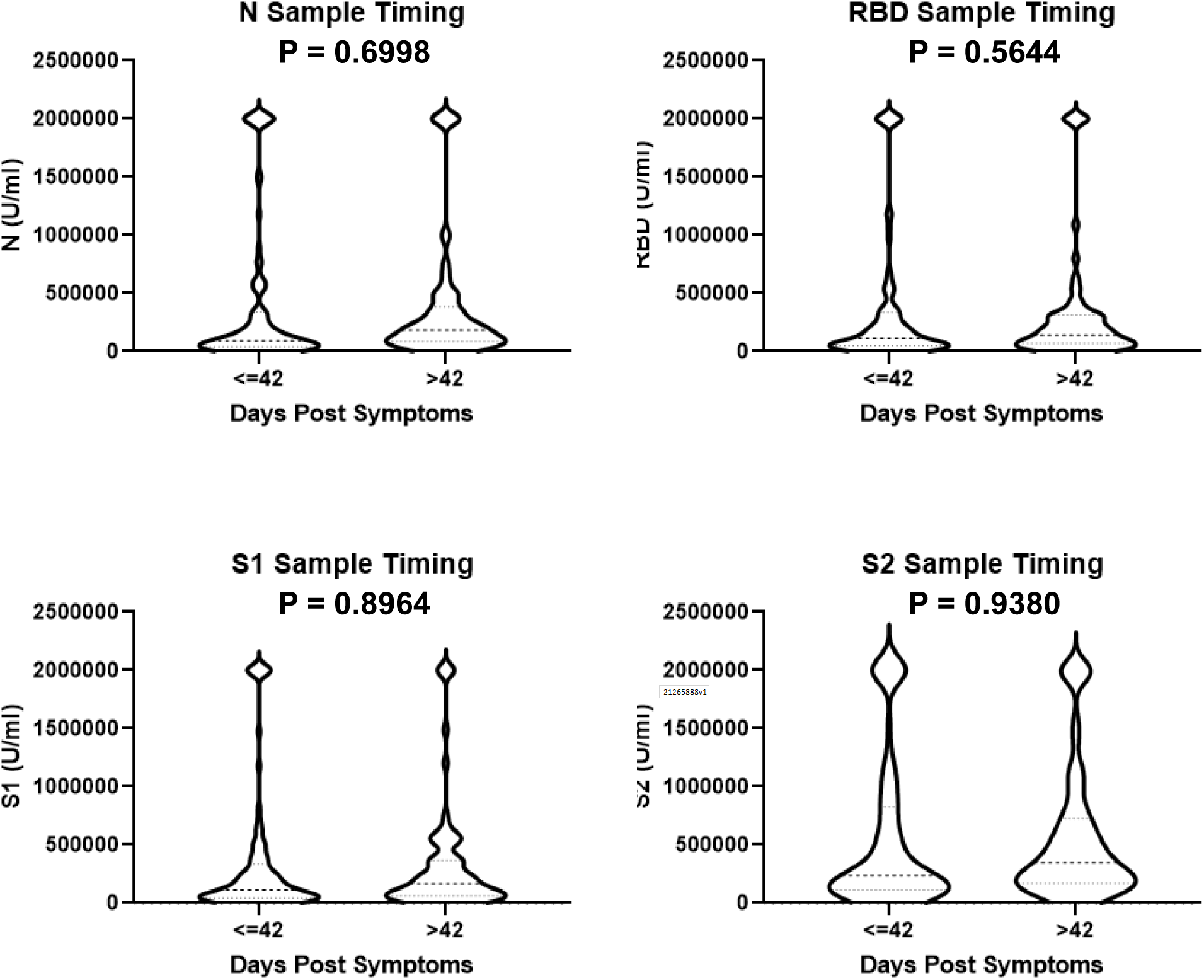
The impact of sample timing on SARS-Cov-2 antibody concentration. Participants were stratified by when blood samples were collected after the onset of COVID-19 symptoms with a split at 42 days (N=97 less than or equal to 42, 75 greater than 42). Antibody concentrations were determined by Bioplex assay and compared using unpaired t-test.

**Figure 6.**
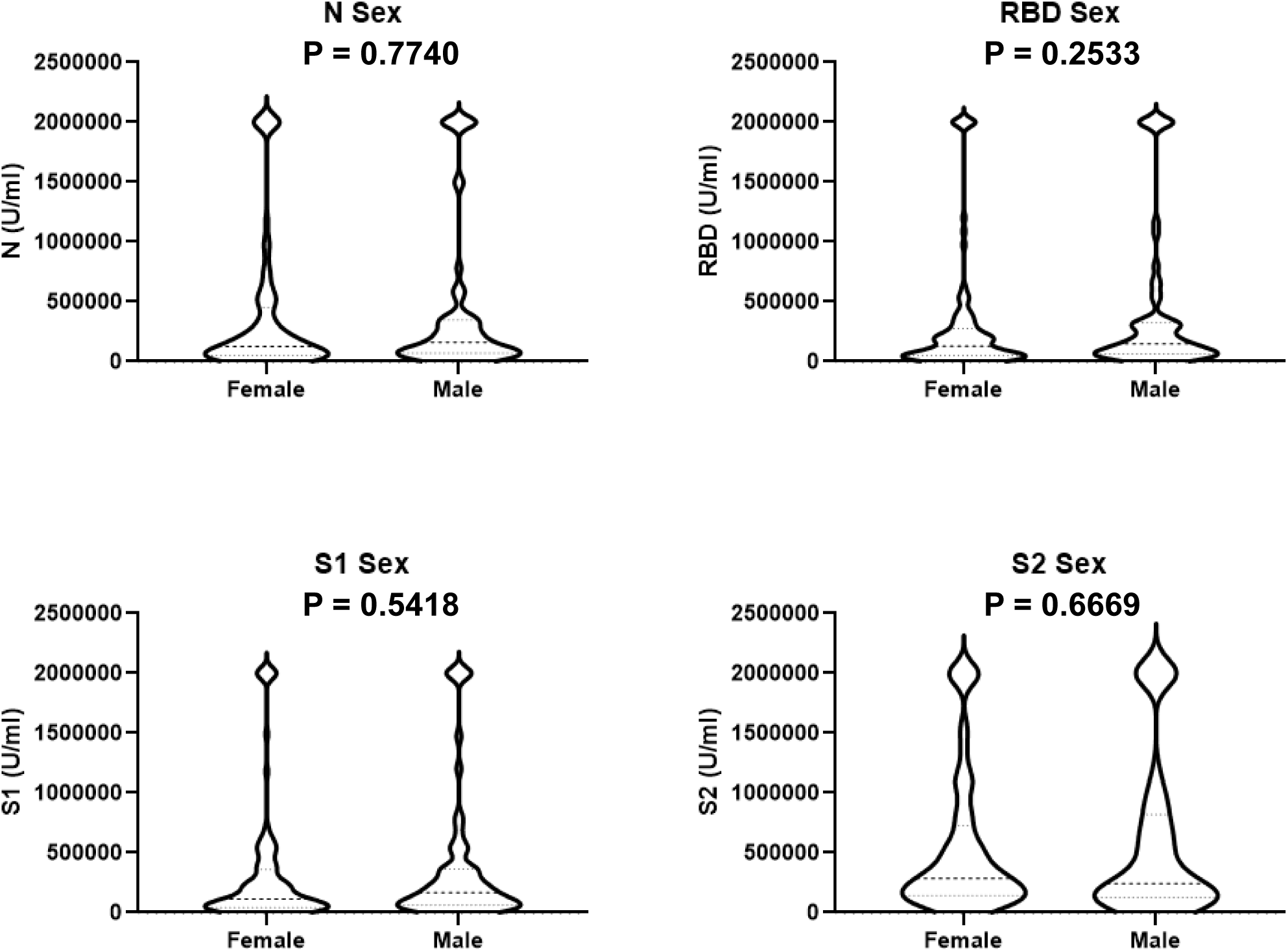
The impact of biologic sex on SARS-Cov-2 antibody concentration. Participants were stratified by sex (N=105 female, 67 male). Antibody concentrations were determined by Bioplex assay and compared using unpaired t-test.

We broke the cohort into 4 groups of under 30, 30-45, 46-59 and over 60 years. When examined by age groups (under 30, 30-45, 46-59 and over 60 years), antibodies against SARS-CoV-2 N, RBD, and S1 were significantly lower in participants under the age of 30 years compared with participants over age 45 years (Figure 7). There was also a trend towards a correlation between older age and serum neutralizing activity. While all participants were recruited as outpatients, we attempted to characterize the severity of disease by measuring the number of days until recovery and if the patient returned to normal activity at the time of convalescent sampling. The mean days to recovery was significantly longer in participants over 45 compared with those in the under 30 and 30-45 groups (Table 2). Similarly, a greater proportion of participants had not yet returned to normal activity in the over 45 group compared with the two young participant groups.

**Table 2.**
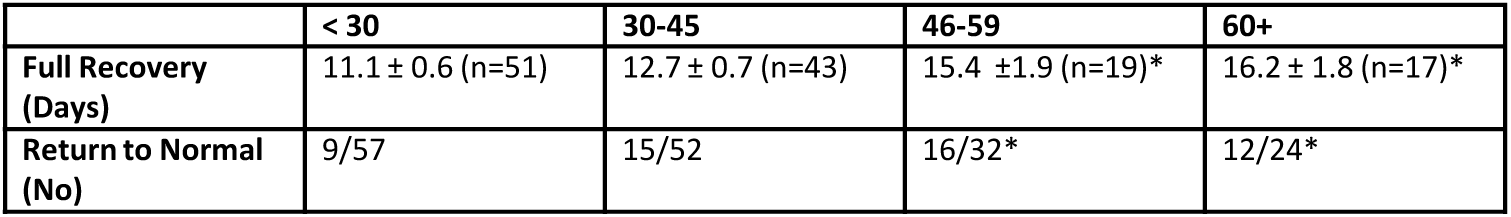

|  | < 30 | 30-45 | 46-59 | 60+ |
| --- | --- | --- | --- | --- |
| Full Recovery (Days) | 11.1 ± 0.6 (n=51) | 12.7 ± 0.7 (n=43) | 15.4 ±1.9 (n=19)* | 16.2 ± 1.8 (n=17)* |
| Return to Normal (No) | 9/57 | 15/52 | 16/32* | 12/24* |

**Figure 7.**
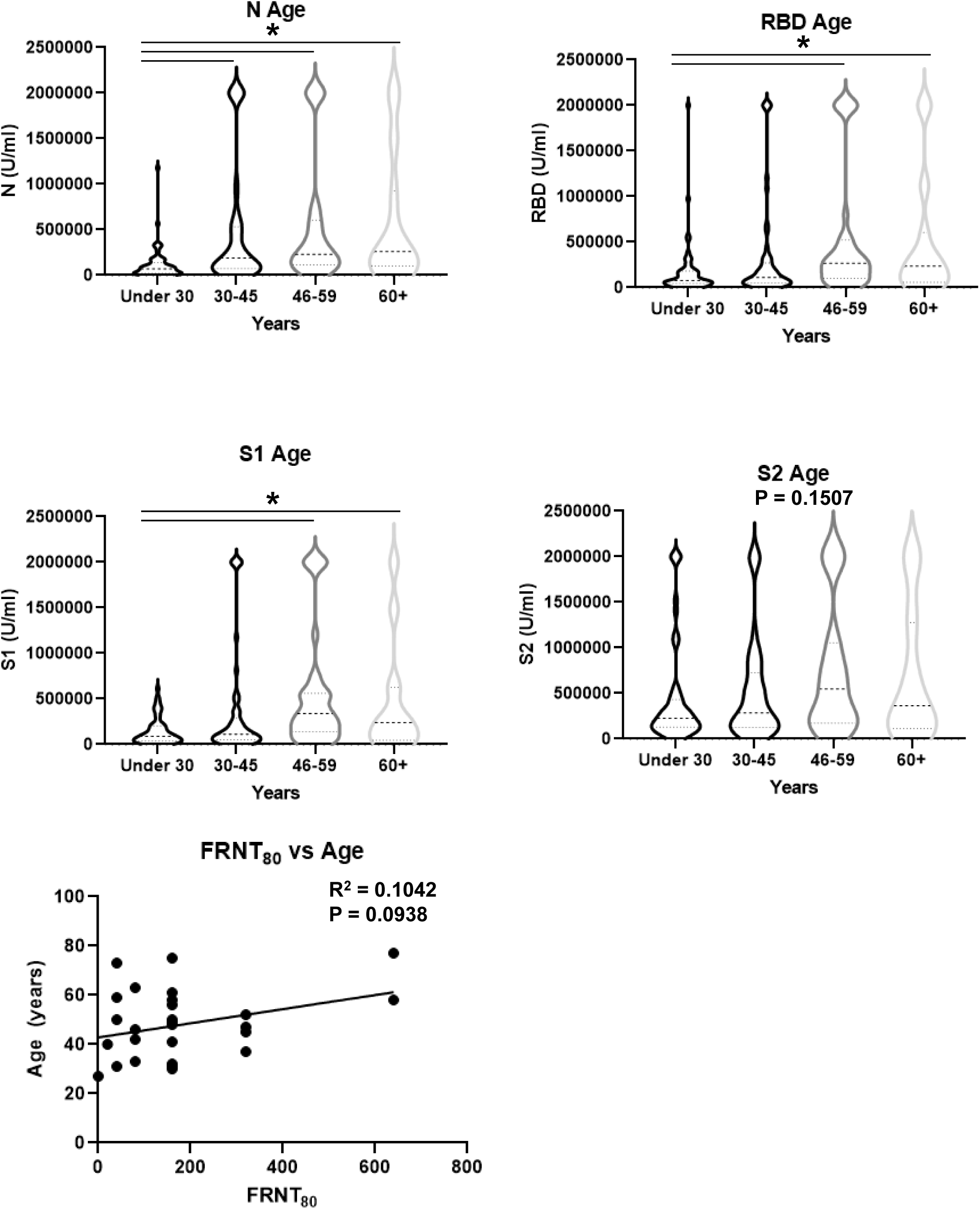
The impact of participant age on SARS-Cov-2 antibody concentration. Participants were stratified by age (N=58 under 30, 55 age 30-45, 36 age 46-59, 24 age 60 and over). Antibody concentrations were determined by Bioplex assay and compared using one-way ANOVA followed by Tukey post-hoc test. Serum neutralization activity and age were compared using simple linear regression.

## Discussion

This study focused on a cohort of COVID-19 outpatients with mild or moderate illness. Many published serology studies have focused on more severe disease, with mild illness limited to a comparator for the robust antibody response in hospitalized participants. More than ten studies have shown that antibody response in severe COVID-19 is greater than in mild cases, as reviewed (11). These findings have been extended by similar observations regarding the presence of neutralizing antibodies in serum (11). While study of severe disease is important, most cases of COVID-19 do not require hospitalization and thus, most individuals infected with SARS-CoV-2 would be expected to have antibody responses similar to those observed in our study. Our finding that 98.8% (171/173) of our outpatient cohort developed measurable antibody against at least one SARS-CoV-2 antigen is consistent with findings in many other studies that suggest that natural infection of variable severity induces humoral immunity against the virus (1).

Given our focus on COVID-19 outpatients, we were able to stratify these patients into groups of low and high responders to perform functional antibody assays and to examine the presence of SARS-CoV-2 specific memory B cells. Microneutralization assays revealed that relatively high and low antibody concentration detected in the Bioplex assay correlated with the amount of functional antibody present. In the low responder group, the highest FRNT_80_ titer observed was 1:80, while nearly all high responders tested were equal to or greater than this level. Surprisingly, two participants had high antibody concentration, but no or low microneutralizing activity, suggesting that antibody level alone may not be protective for some patients. Correlation of microneutralization titer with antibody concentration revealed a positive relationship between high spike protein antibody and functional ability, as one would predict given that spike is the target of neutralizing antibodies. The correlation between neutralizing activity and N protein antibody was less strong. Others have shown that SARS-CoV-2 specific memory IgG producing B cells are formed in both mild and severe patients (12, 13). In severe COVID-19 patients, memory B cells were found in all patients studied, however only 80% had neutralizing antibody. In our study, all antibody high responders had detectable spike-specific memory B cells, although the number of memory B cells did not correlate with antibody concentration. All participants tested for memory B cells had serum neutralizing activity of variable titer. Spike-specific B cell number trended towards correlation with microneutralization titer. These data suggest that high responding outpatients form memory B cells which may provide longer lasting protection against SARS-CoV-2 than serum antibody concentration would indicate.

By measuring IgG against 4 four SARS-CoV-2 antigens, we were able to characterize the relative immunodominance of the different antigens. Antibody response to N protein only partially correlates with spike protein antibody responses. Our data showed that some participants responded to N better than to spike proteins and vice versa, with the relationship between N and S1 antibodies being the strongest positive correlation. S1 and S2 antibody response was positively correlated, but not entirely linear suggesting differential immune recognition of these spike protein domains among the cohort. Interestingly, the incidence of a low antibody response was lowest for the S2 domain of the spike protein when compared with S1, RBD, or N proteins. In the rare occurrence of very low antibody response to 1 or more antigens tested, females were more likely than males to display this poor response phenotype.

Our study did not show an impact of sample timing or biologic sex on serum antibody concentrations. Numerous studies have shown a degree of antibody waning 2 to 3 months after infection. Our cohort spanned 1 to 4 months after the onset of symptoms with a median near 42 days. In outpatients, we did not observe a decrease in serum antibody in this time period. Several studies have shown no effect of biologic sex on serum antibody titer, however neutralizing antibody levels have been shown to correlate with male sex, as reviewed (11, 14). This is consistent with our findings of antibody concentration; however, we did not see a significant difference in neutralizing antibody titer and sex (data not shown). We saw a trend towards more spike-specific B cells in males versus females. The effect of sex on SARS-CoV-2 antibody response does not appear to be a primary driver of response in outpatients in this study.

Importantly, the largest predictor of antibody response in our study was participant age. Participants under the age of 30 had significantly lower antibody concentration for N, S1, and RBD proteins compared with older participants. This is consistent with some studies that have shown a correlation between IgG levels and age, with lower antibody concentration in younger adults (14, 15). In those studies, conflicting results were found regarding neutralizing antibody titers as one study found an increase in older adults, while the other showed higher levels in younger adults. Other research did not show a relationship between age and antibody concentration, as reviewed (11). In our study, we observed a trend towards a greater number of spike-specific memory and IgG-producing B cells in younger participants versus older adults, which may reflect the lack of capacity of naïve B cells in older participants. Regardless, serum antibody concentration was lower in those under the age of 30. Neutralizing activity in serum trended to increase with age, but was not significant in our study. One possible explanation for the age effect observed would be the severity of disease in our cohort. To address this issue, we examined the number of days until recovery and if a return to normal was seen at the time of sampling. We did show that recovery and a lack of return to normal activities was significantly longer in participants over 45 years, compared with younger participants. However, these parameters were not different in participants under 30 or those from 30-45, while antibody concentration was lower between these two groups. These data suggest that measures of outpatient severity do not likely explain the lower antibody response in young participants in our study. It is possible that younger participants had better controlled viral replication, and thus, there was less viral antigen to stimulate ongoing immune responses.

In summation, we characterized serum antibody and B cell responses to SARS-CoV-2 in a cohort of COVID-19 outpatients. We did so using well validated assays to generate reliable antibody concentration, microneutralization, and flow cytometry data. The primary finding was that participant age affected serum antibody concentration, with younger participants producing lower antibody responses. We also showed that antibody concentration was predictive of microneutralization activity, but not the number of spike-specific B cells in blood. In whole, these data inform current vaccination policy as young previously infected people likely require vaccine boosting. Further, antibody concentration may not directly relate to the presence of SARS-CoV-2 specific memory B cells capable of providing protection against infection and/or severity of disease. As serum antibody levels wane following infection, memory B cells may persist longer to provide sustained protection. These questions will need to be answered at later time points following infection.

## Methods

### Cohort

All participant samples were collected as part of the US Influenza Vaccine Effectiveness (FLUVE) Network – Pittsburgh site. This network is an annual prospective study of outpatients who seek care for acute respiratory illness. This study was approved by the University of Pittsburgh Institutional Review Board. All 173 participants were adults (≥ 18 years of age) and were SARS-CoV-2 positive by molecular testing. Relevant participant information was subsequently extracted from the medical record. One participant did not disclose sex and another participant did not have the timing of blood sampling relative to illness recorded.

### Blood Processing

Blood was collected in Becton Dickinson SST (serum) or CPT (peripheral blood mononuclear cells (PBMC)) and processed following manufacturer’s instructions. Serum was aliquoted and stored at −80° C prior to analyses. PBMC were aliquoted, frozen, and stored in liquid nitrogen prior to analyses.

### Antibody Assay

SARS-CoV-2 specific antibody concentrations were determined using a commercially available Biorad Bioplex Kit; Bio-Plex Pro Human IgG SARS-CoV-2 N/RBD/S1/S2 4-Plex Panel (catalog #12014634). Analyses of serum samples were performed by manufacturer’s instructions. Banked pre-COVID-19 serum samples were utilized as negative controls. SARS-CoV-2 antibody standards were provided by Biorad, with concentrations of U/ml. To control for plate-to-plate variability, five samples were repeated on all three assay plates run. Concordance was very high between assay runs (r^2^ of 0.85-0.96), suggesting that combining data was appropriate (Supplemental Figure 3). In addition, World Health Organization SARS-CoV-2 antibody reference samples were assayed to compare with values obtained using the Bioplex kit. The COVID-19 convalescent plasma panel (NIBSC 20/118) was obtained from the National Institute for Biological Standards and Control, United Kingdom. Concentrations derived using the Bioplex kit correlated with the reported WHO reference sample concentrations (Supplemental Figure 4).

### Neutralization Assay

Antibody neutralization of SARS-CoV-2 was performed as published (16). Spots in Vero E6 cell cultures were imaged, counted, and processed using an ImmunoSpot Counter (CTL). Foci were counted in experimental wells and compared to control wells. The dilution of serum at which 80% of foci are neutralized is reported as the FRNT_80_.

### B Cell Flow Cytometry

Frozen PBMCs were rapidly thawed in a water bath at 37°C, rinsed with 1ml flow sorter buffer (1x phosphate buffered saline, 2% newborn calf serum, and 0.1% sodium azide) and centrifuged at 500 x g for 5 minutes, 4°C. Cells were resuspended in 100 μl of sorter buffer with 2.5 μg of human Fc Block (clone Fc.3216) for 10 minutes at room temperature. Next, cells were incubated with 1 mM PE-Cy5 decoy for 5 minutes at room temperature followed by 1 mM SARSCoV2 Spike-PE B cell tetramer for 25 minutes on ice in the dark using decoy and tetramer created in the lab as previously described (13, 17). Cells were then washed and incubated with anti-PE magnetic beads (Miltenyi) and passed over a LS column on a quadroMACS magnet. Eluted cells were stained for 30 minutes at 4° C in the dark with the following anti-human antibodies (clone): CD19 (SJ25C1), CD3 (UCHT1), CD14 (HCD14), CD16 (B73.1), CD21 (Bu32), CD27 (M-T271), IgD (IA6-2), IgM (MHM-88) and IgG (G18-145). Next, cells were washed with 1 ml flow sorter buffer, centrifuged at 500 x g for 5 minutes, and fixed with 200 μl 2% paraformaldehyde at 4° C for 10 minutes. Cells were washed and resuspended in flow sorter buffer and collected on an LSRFortessa (Beckton-Dickson). Data was analyzed using FlowJo software (TreeStar, Ashland, OR).

### Statistics

Statistical analyses were performed using GraphPad Prism Software (San Diego, CA). Correlation data was analyzed by simple linear regression with an alpha of 0.05 for significance. Differences between two means were analyzed by unpaired t-test with an alpha of 0.05 for significance. Differences between more than two means were analyzed by one-way ANOVA followed by Tukey post-hoc test with an alpha of 0.05 for significance. Cohort data sets are shown as violin plots with the median and quartiles indicated. Alternatively, data sets are displayed as dot plots with mean and standard error of the mean displayed.

## Data Availability

All data produced in the present study are available upon reasonable request to the authors.

## References

1. Cromer D, Juno JA, Khoury D, Reynaldi A, Wheatley AK, Kent SJ, and Davenport MP. Prospects for durable immune control of SARS-CoV-2 and prevention of reinfection. Nat Rev Immunol. 2021;21(6):395–404.

2. Gonzalez F, Zepeda O, Toval-Ruiz C, Matute A, Vanegas H, Munguia N, Centeno E, Reyes Y, Svensson L, Nordgren J, et al. Antibody response to SARS-CoV-2 infection over six months among Nicaraguan outpatients. medRxiv. 2021.

3. Grossberg AN, Koza LA, Ledreux A, Prusmack C, Krishnamurthy HK, Jayaraman V, Granholm AC, and Linseman DA. A multiplex chemiluminescent immunoassay for serological profiling of COVID-19-positive symptomatic and asymptomatic patients. Nat Commun. 2021;12(1):740.

4. Peghin M, De Martino M, Fabris M, Palese A, Visintini E, Graziano E, Gerussi V, Bontempo G, D’Aurizio F, Biasotto A, et al. The Fall in Antibody Response to SARS-CoV-2: a Longitudinal Study of Asymptomatic to Critically Ill Patients Up to 10 Months after Recovery. J Clin Microbiol. 2021;59(11):e0113821.

5. Roltgen K, Powell AE, Wirz OF, Stevens BA, Hogan CA, Najeeb J, Hunter M, Wang H, Sahoo MK, Huang C, et al. Defining the features and duration of antibody responses to SARS-CoV-2 infection associated with disease severity and outcome. Sci Immunol. 2020;5(54).

6. Sasson JM, Campo JJ, Carpenter RM, Young MK, Randall AZ, Trappl-Kimmons K, Oberai A, Hung C, Edgar J, Teng AA, et al. Diverse Humoral Immune Responses in Younger and Older Adult COVID-19 Patients. mBio. 2021;12(3):e0122921.

7. Stromer A, Rose R, Grobe O, Neumann F, Fickenscher H, Lorentz T, and Krumbholz A. Kinetics of Nucleo- and Spike Protein-Specific Immunoglobulin G and of Virus-Neutralizing Antibodies after SARS-CoV-2 Infection. Microorganisms. 2020;8(10).

8. Wellinghausen N, Voss M, Ivanova R, and Deininger S. Evaluation of the SARS-CoV-2-IgG response in outpatients by five commercial immunoassays. GMS Infect Dis. 2020;8(Doc22.

9. Anichini G, Terrosi C, Gandolfo C, Gori Savellini G, Fabrizi S, Miceli GB, and Cusi MG. SARS-CoV-2 Antibody Response in Persons with Past Natural Infection. N Engl J Med. 2021;385(1):90–2.

10. Long QX, Liu BZ, Deng HJ, Wu GC, Deng K, Chen YK, Liao P, Qiu JF, Lin Y, Cai XF, et al. Antibody responses to SARS-CoV-2 in patients with COVID-19. Nat Med. 2020;26(6):845–8.

11. Post N, Eddy D, Huntley C, van Schalkwyk MCI, Shrotri M, Leeman D, Rigby S, Williams SV, Bermingham WH, Kellam P, et al. Antibody response to SARS-CoV-2 infection in humans: A systematic review. PLoS One. 2020;15(12):e0244126.

12. Varnaite R, Garcia M, Glans H, Maleki KT, Sandberg JT, Tynell J, Christ W, Lagerqvist N, Asgeirsson H, Ljunggren HG, et al. Expansion of SARS-CoV-2-Specific Antibody-Secreting Cells and Generation of Neutralizing Antibodies in Hospitalized COVID-19 Patients. J Immunol. 2020;205(9):2437–46.

13. Rodda LB, Netland J, Shehata L, Pruner KB, Morawski PA, Thouvenel CD, Takehara KK, Eggenberger J, Hemann EA, Waterman HR, et al. Functional SARS-CoV-2-Specific Immune Memory Persists after Mild COVID-19. Cell. 2021;184(1):169–83 e17.

14. Chvatal-Medina M, Mendez-Cortina Y, Patino PJ, Velilla PA, and Rugeles MT. Antibody Responses in COVID-19: A Review. Front Immunol. 2021;12(633184.

15. Yang HS, Costa V, Racine-Brzostek SE, Acker KP, Yee J, Chen Z, Karbaschi M, Zuk R, Rand S, Sukhu A, et al. Association of Age With SARS-CoV-2 Antibody Response. JAMA Netw Open. 2021;4(3):e214302.

16. Xu L, Doyle J, Barbeau DJ, Le Sage V, Wells A, Duprex WP, Shurin MR, Wheeler SE, and McElroy AK. A Cross-Sectional Study of SARS-CoV-2 Seroprevalence between Fall 2020 and February 2021 in Allegheny County, Western Pennsylvania, USA. Pathogens. 2021;10(6).

17. Taylor JJ, Martinez RJ, Titcombe PJ, Barsness LO, Thomas SR, Zhang N, Katzman SD, Jenkins MK, and Mueller DL. Deletion and anergy of polyclonal B cells specific for ubiquitous membrane-bound self-antigen. J Exp Med. 2012;209(11):2065–77.

