## Supplemental Material for "SARS-CoV-2 Antibody Response is Associated with Age in Convalescent Outpatients"

**Supplemental Figure Legends**

Supplemental Figure 1 – Comparison of microneutralization titers and IgG concentrations for each antigen tested. Bioplex derived antibody concentration (U/ml) were compared with microneutralization titer (1:X) by simple linear regression (N=56).

Supplemental Figure 2 – Memory B cell flow cytometry gating strategy. Lymphocytes were gated by CD19 positivity, CD21/CD27 status, and IgD, IgM, and IgG positivity to identify spike specific memory B cells, isotype switched B cells, and IgG producing B cells. Spike specificity was determined using a spike tetramer (N=21 high responders).

Supplemental Figure 3 – Inter-assay Bioplex plate concordance comparison. All study samples were run on a total of 3 Bioplex plates. A set of four samples were repeatedly run on all three plates and concentration values for each antigen were compared between plates using simple linear regression (N=16 antibody values).

Supplemental Figure 4 – Comparison of Bioplex and ELISA concentrations for WHO reference samples. WHO reference samples (COVID-19 convalescent plasma panel NIBSC 20/118) were obtained from the National Institute for Biological Standards and Control, United Kingdom. IgG concentration (U/ml) from Bioplex were compared with values derived by ELISA. Graphs are nonlinear, sigmoidal 4PL least squares fit.

Supplemental Figure 1

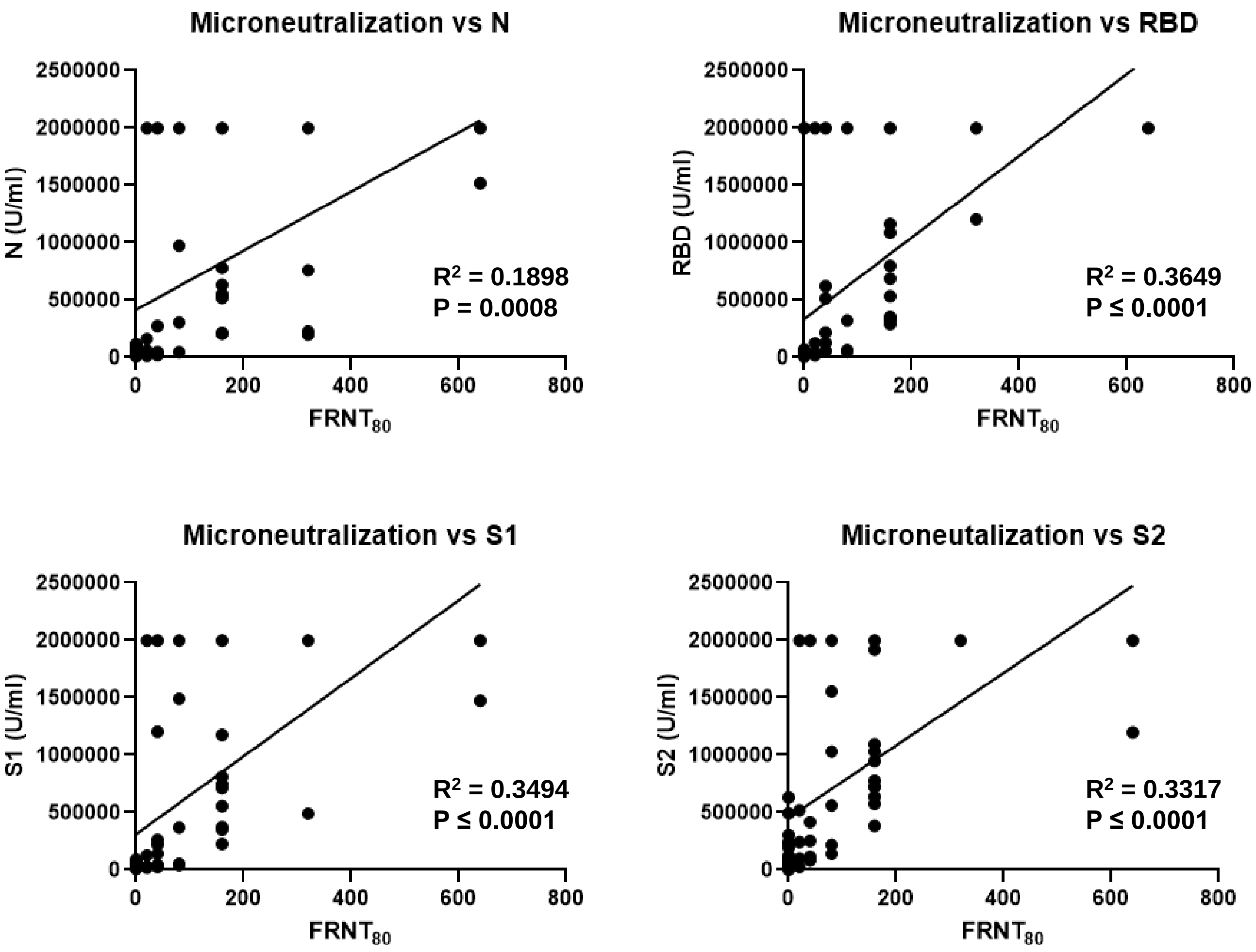

### Supplemental Figure 2

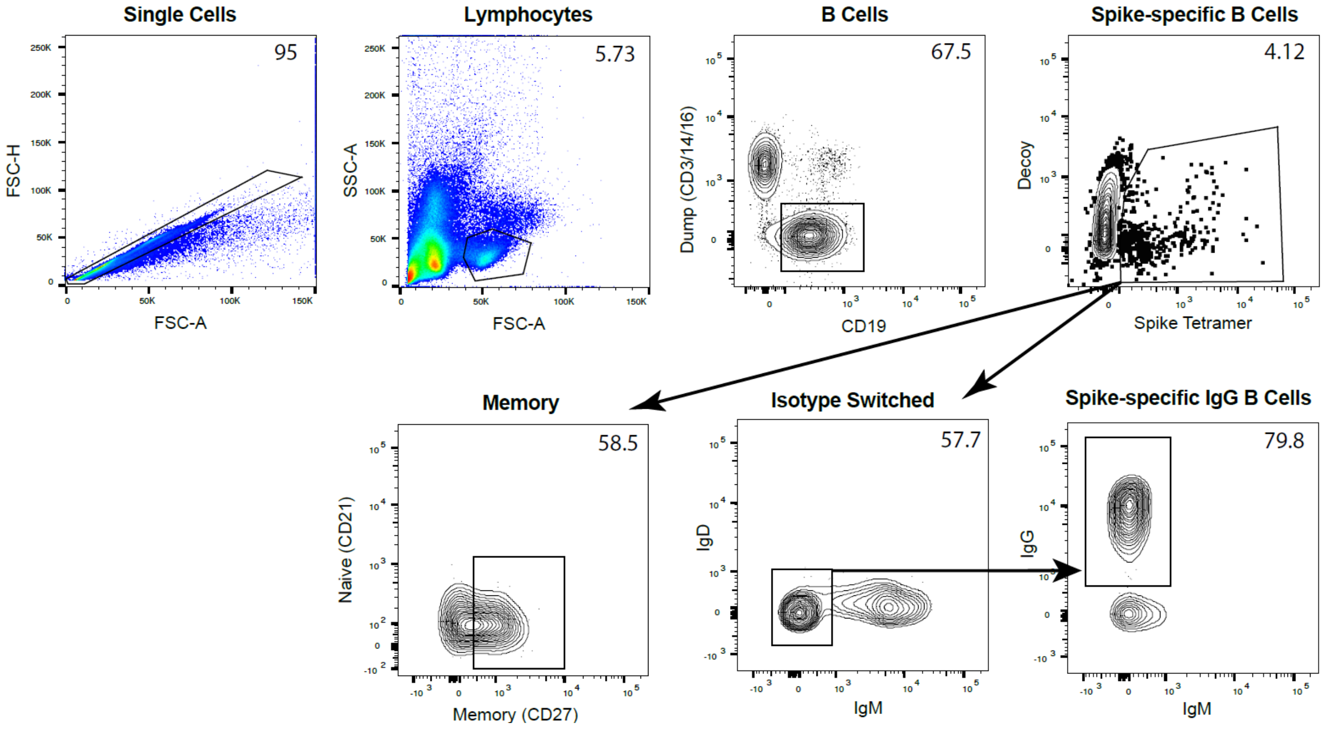

### Supplemental Figure 3

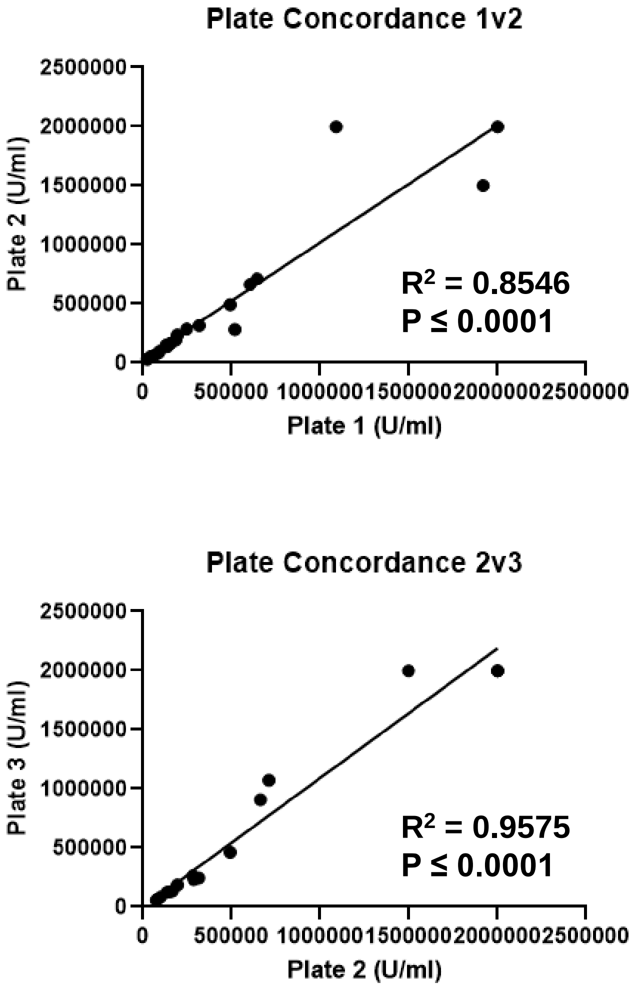

Supplemental Figure 4

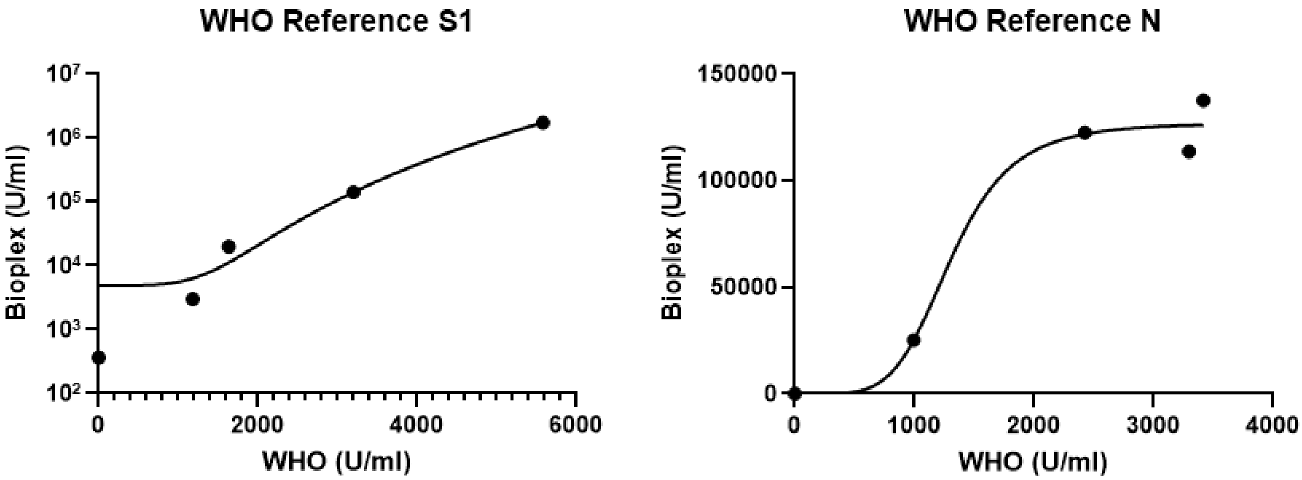
